# A Causal Multi-modal AI Model Stratifies Residual Risk and Identifies Candidates for Treatment Escalation in Node-Positive HR+/HER2− Early Breast Cancer

**DOI:** 10.64898/2026.09.15.26362492

**Authors:** Thomas Bachelot, Sylvie Chabaud, Jérôme Lemonnier, Paul H Cottu, Florence Dalenc, Frederick M. Howard, Lajos Pusztai, Cerise Tang, Dhruva Biswas, Ken Zeng, Jan Witowski, Krzysztof J Geras, Fabrice André, Frédérique Madeleine Penault-Llorca

## Abstract

**PURPOSE:** Prognostic biomarkers that estimate residual recurrence risk after standard adjuvant therapy to guide further treatment escalation are an unmet clinical need. This study used Ataraxis Breast CTX, a causal multi-modal AI model integrating clinical variables and histopathology, to stratify residual risk in clinically defined high-risk patients. We aimed to identify patients who have excellent outcomes on standard-of-care chemoendocrine therapy and patients who may benefit from additional therapies such as everolimus or CDK4/6 inhibitors.

**METHODS:** CTX combines clinical variables with image features from H&E images to produce estimates of recurrence risk (CTX-prognostic) and chemotherapy benefit (CTX-benefit). Prospective-retrospective validation was performed in the phase III UNIRAD trial to assess CTX-prognostic’s performance in the subset of patients who had not received neoadjuvant chemotherapy, who had received adjuvant chemotherapy, and who had H&E slides available (n = 556). The primary endpoint in this study was disease-free survival (DFS). As an exploratory analysis, we evaluated CTX-benefit’s ability to predict benefit from adjuvant everolimus.

**RESULTS:** CTX-prognostic effectively stratified patients into high- and low-risk groups. Across all patients, 5-year DFS was significantly higher in the low-risk group (93%) than in the high-risk group (80%). Additionally, treatment benefit from everolimus differed significantly by CTX-benefit score in an exploratory multivariable analysis adjusting for age, tumor size, nodal status, grade, and menopausal status (interaction p = 0.01).

**CONCLUSION:** CTX identified patients with good outcomes under chemoendocrine therapy alone and patients with residual risk who may benefit from treatment escalation. Additionally, CTX was found to be predictive of everolimus benefit. These results suggest that CTX may effectively stratify clinically high-risk HR+/HER2− patients by residual risk and may inform selection of adjuvant escalation strategies.

## INTRODUCTION

Patients diagnosed with early-stage hormone receptor-positive (HR+), human epidermal growth factor receptor 2-negative (HER2-) breast cancer are typically treated with surgery followed by endocrine therapy for 5-10 years.^1^ Patients with lymph node-positive disease experience higher rates of recurrence, often warranting the addition of adjuvant chemotherapy.^2,3^ However, despite chemoendocrine therapy, 20% of patients with pN1 breast cancer and 34% of patients with pN2 breast cancer still recur.^4^ This has led to the development of new targeted therapies such as CDK4/6 and mTOR inhibitors.^5–8^ The phase III UNIRAD and SWOG 1207 trials tested the efficacy of the mTOR inhibitor everolimus as adjuvant therapy in patients with high-risk, HR+/HER2−, node-positive stage II-III breast cancer.^7,8^ Neither of these trials demonstrated a significant difference in disease-free survival between the control and everolimus arms.^7,8^

Precision medicine tests have provided physicians with prognostic information beyond traditional staging criteria, thereby allowing more individualized therapy decision-making. Specifically, gene expression profiling tests, such as the commercially available 12-, 21-, and 70-gene assays, identify patients who may avoid adjuvant chemotherapy with no reduction in recurrence-free survival.^9–11^ However, these assays have not been validated for adjuvant therapy decision-making beyond chemotherapy de-escalation. Thus, patient selection for additional therapies continues to rely on clinicopathologic risk criteria, an approach that is suboptimal, as illustrated by the modest treatment effects observed in escalation trials.^5,6^ The toxicities and financial burden associated with adjuvant CDK4/6 inhibitor therapy motivate the search for improved risk stratification. Importantly, while nodal involvement confers elevated recurrence risk, outcomes remain heterogeneous, and current clinicopathologic criteria cannot reliably identify patients with sufficiently low residual risk to forgo further escalation.

Here, we evaluate Ataraxis Breast CTX, a causal multi-modal artificial intelligence (AI) model for chemotherapy benefit, in an analysis of the UNIRAD trial. CTX integrates digital histopathology features extracted from H&E-stained tumor slides with clinical features to estimate the probability of remaining recurrence-free and chemotherapy benefit (see *Methods*).^12^ We hypothesize that CTX may identify clinically high-risk patients (the population represented in the UNIRAD trial) who would experience excellent outcomes with standard-of-care chemoendocrine therapy. Additionally, we hypothesize that individuals with high residual risk who are unlikely to benefit from chemotherapy are more likely to derive benefit from escalation therapies such as everolimus, evaluated here, or CDK4/6 inhibitors.

## METHODS

### Development of CTX

CTX is a causal multi-modal AI model that integrates pathology features with clinical variables (**Figure 1a**). CTX was developed using a metalearner framework for estimating heterogeneous treatment effects,^13^ which enables it to explicitly make counterfactual predictions assuming different therapies. CTX generates these predictions from a multi-stage pipeline that converts raw slides into patient-level representations. First, whole-slide images are tiled into patches that are next encoded into features using a pathology foundation model.^14^ These pathology features are then fused with clinical variables to generate patient-level representations, which are used to estimate, for each patient, the probability of remaining recurrence-free under 1) endocrine therapy only or 2) chemoendocrine therapy. Next, these scores undergo post-hoc calibration to generate the final predictions.^15^ These calibrated predictions yield two scores. CTX-prognostic is the predicted 5-year recurrence risk under chemoendocrine therapy, while CTX-benefit is the predicted chemotherapy benefit, defined as the difference between recurrence risk on endocrine vs chemoendocrine therapy. **Figure S1** presents a patient-level scatterplot of CTX-prognostic against CTX-benefit in the UNIRAD data, illustrating the relationship between predicted recurrence risk and predicted chemotherapy benefit (Pearson’s *r* = 0.77, p < 0.001). CTX was trained on 9,141 patients from multiple independent institutions, and was locked at the end of the development phase. Notably, no data from the UNIRAD trial were used for model development, and there was no subsequent retraining or recalibration for this validation study (**Figure 1b**).

**FIG 1.**
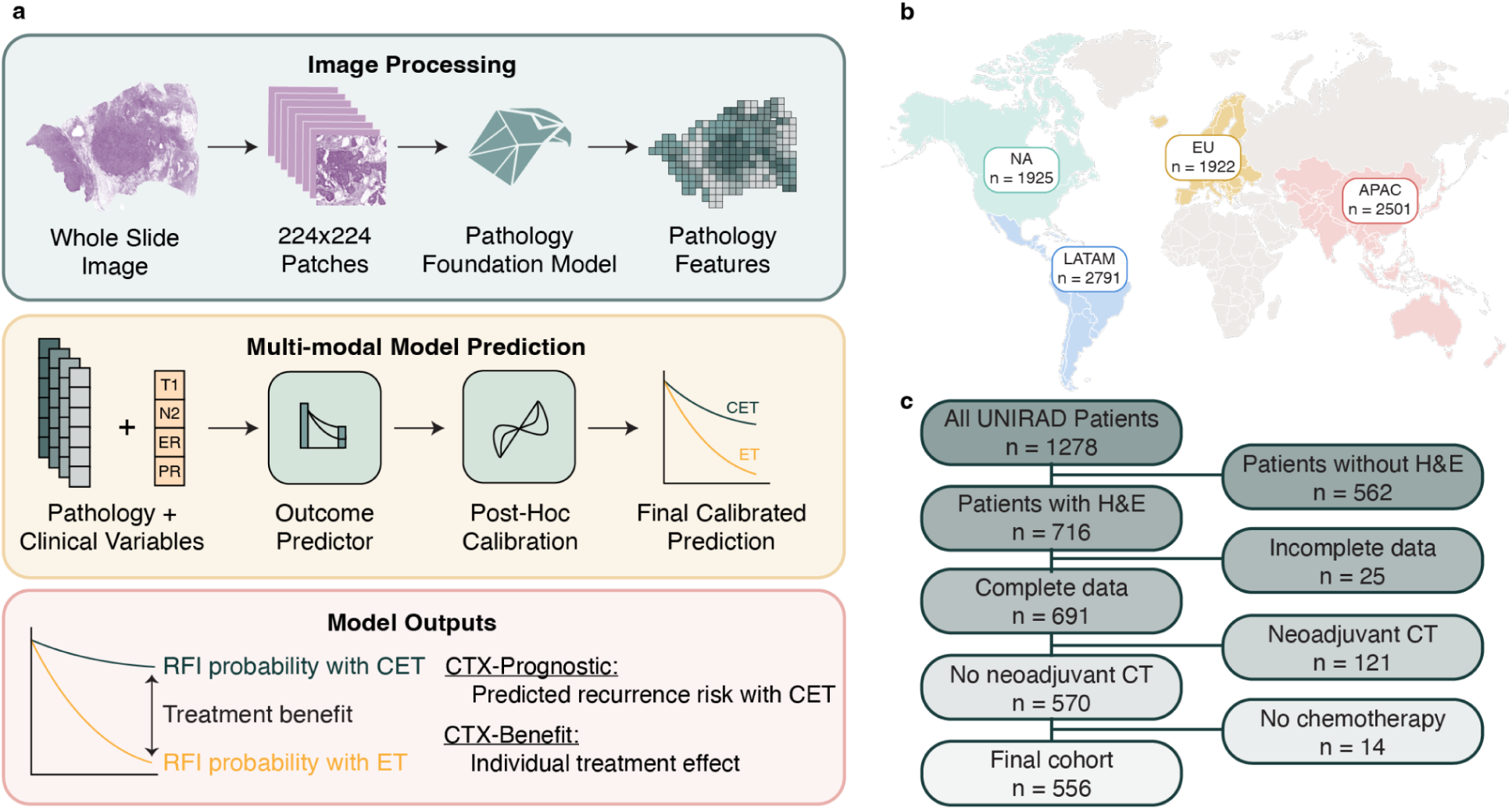
Overview of CTX training and score generation. a) Overview of the CTX model. Whole-slide images are split into patches and encoded into patch-level embeddings using a pathology foundation model. Patch-level embeddings are then aggregated into a slide-level representation. Slide-level embeddings were integrated with clinical variables to generate a multi-modal representation, which was used to predict recurrence-free interval under endocrine therapy only and chemoendocrine therapy. These predictions then undergo post-hoc calibration. From these calibrated predictions, we derive CTX-prognostic (predicted risk under adjuvant chemoendocrine therapy) and CTX-benefit (predicted chemotherapy benefit). b) The training dataset (n = 9,141) comprised patients from North America (NA, n = 1,925), Europe (EU, n = 1,922), Asia-Pacific (APAC, n = 2,501), and Latin America (LATAM, n = 2,791). c) Flowchart of patient selection starting with all patients enrolled in the phase III UNIRAD trial (n = 1,278), narrowed by patients with a H&E-stained slide available for imaging (n = 716, 56%). Patients who did not have complete clinical data (n = 25), who received neoadjuvant chemotherapy (n = 121), or did not receive adjuvant chemotherapy (n = 14) were removed from the study cohort, leaving 556 patients in the final analysis group. ET denotes endocrine therapy only; CET, chemoendocrine therapy; CT, chemotherapy; RFI, recurrence-free interval.

### Selection of Binarization Cut-Offs

CTX generates continuous, calibrated probabilities of recurrence, and analyses use these scores in continuous form whenever possible as they are the best reflection of the model’s output. However, continuous probabilities are not directly actionable as treatment decisions are binary, and Kaplan-Meier (KM) estimates and categorical statistics are conventionally reported by risk groups. Therefore, we applied pre-defined thresholds for recurrence risk and chemotherapy benefit.

The low- versus high-risk threshold was selected using three external datasets not used for training CTX (n = 1,150). The goal was to identify a low-risk group with sufficiently low recurrence risk to forgo treatment escalation. Low-risk was defined as a KM-estimated 10-year DFS ≥ 95%, a bar higher than the outcomes reported in both arms of the NATALEE and monarchE trials.^5,6^ To find the appropriate CTX-prognostic cutoff, we tested 1,000 candidate thresholds evenly spaced across the observed score range. Across 1,000 bootstrap resamples, we selected, within each resample, the highest threshold at which low-risk patients met the ≥ 95% DFS criterion. The final threshold was the median of the 1,000 thresholds (6.5%). Patients in the UNIRAD cohort were subsequently stratified into high- and low-risk groups using this pre-specified threshold. Separately, a threshold of 2% predicted chemotherapy benefit, which has been used in previous studies of CTX,^12^ was used to classify UNIRAD patients into high chemotherapy benefit vs low chemotherapy benefit groups.

### Patients and Study Design

Between June 2013 and March 2020, 1,278 women aged 18 years or older with ER+/HER2− early breast cancer at high risk of relapse were enrolled in the UNIRAD trial (**Figure 1c**). Selection criteria for UNIRAD were: high nodal involvement (≥ 4 positive lymph nodes) and/or persistent nodal involvement after neoadjuvant chemotherapy (≥ 1 positive lymph node), or 1-3 positive lymph nodes at primary surgery and an EndoPredict (EPclin) score ≥ 3.3.^11^ For our analysis, of the 1,278 patients enrolled in the UNIRAD trial, 562 did not have an H&E slide available, 121 patients received neoadjuvant chemotherapy, 25 did not have clinical data, and 14 patients did not receive adjuvant chemotherapy. Patients who received neoadjuvant treatment were excluded because prior cytotoxic exposure may alter tumor biology and image features, potentially confounding treatment effect estimates from an assay developed on treatment-naive samples. Patients without adjuvant chemotherapy were excluded because this analysis aimed to determine whether everolimus confers additional benefit alongside chemotherapy. The final analysis cohort included 556 patients with characteristics described in **Table 1**. H&E slides were collected from each participating site and digitized centrally by UNICANCER using a Pannoramic Scan II (3DHISTECH) scanner.

**TABLE 1:**
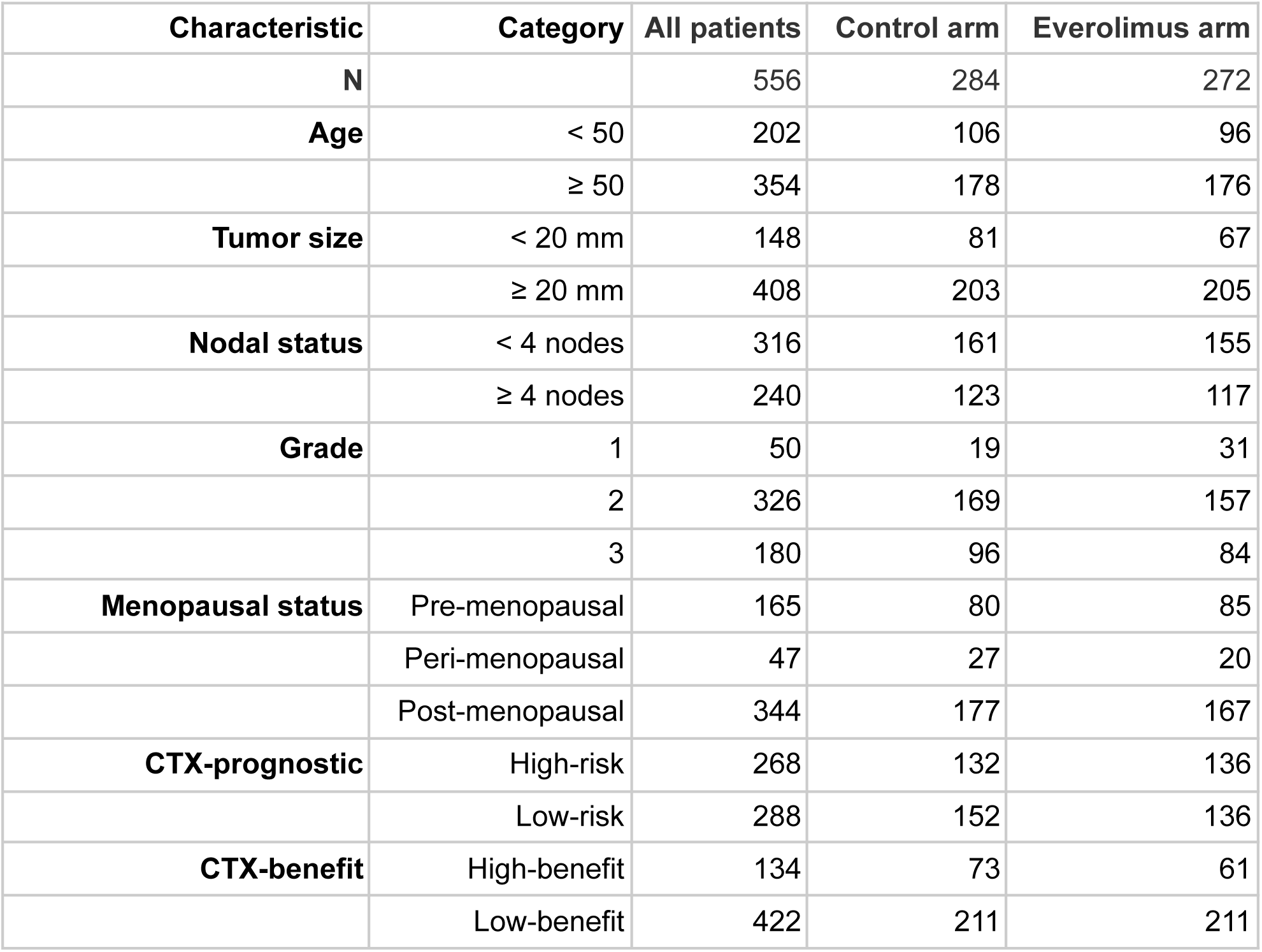
Patient Characteristics by trial arm. No significant differences in clinical characteristics between treatment arms were detected (all chi-square p > 0.05).

| Characteristic | Category | All patients | Control arm | Everolimus arm |
| --- | --- | --- | --- | --- |
| **N** |  | 556 | 284 | 272 |
| **Age** | < 50 | 202 | 106 | 96 |
|  | ≥ 50 | 354 | 178 | 176 |
| **Tumor size** | < 20 mm | 148 | 81 | 67 |
|  | ≥ 20 mm | 408 | 203 | 205 |
| **Nodal status** | < 4 nodes | 316 | 161 | 155 |
|  | ≥ 4 nodes | 240 | 123 | 117 |
| **Grade** | 1 | 50 | 19 | 31 |
|  | 2 | 326 | 169 | 157 |
|  | 3 | 180 | 96 | 84 |
| **Menopausal status** | Pre-menopausal | 165 | 80 | 85 |
|  | Peri-menopausal | 47 | 27 | 20 |
|  | Post-menopausal | 344 | 177 | 167 |
| **CTX-prognostic** | High-risk | 268 | 132 | 136 |
|  | Low-risk | 288 | 152 | 136 |
| **CTX-benefit** | High-benefit | 134 | 73 | 61 |
|  | Low-benefit | 422 | 211 | 211 |

### Statistical Analysis

The primary endpoint in this analysis was DFS, measured from date of randomization to first recurrence of breast cancer or death from any cause. Secondary endpoints were also evaluated: distant metastasis-free survival (DMFS), defined as the time from randomization to distant metastasis or death, and overall survival (OS). All endpoints were pre-specified endpoints of the UNIRAD trial.^7^ To evaluate discrimination of CTX-prognostic, we used C-index.^16–18^ The KM estimator was used to estimate the survival curves, from which 5-year DFS rates were derived. A two-sided log-rank test was used to compare KM curves. A multivariable Cox proportional hazards model was fitted with CTX-prognostic score as a continuous variable, adjusted for patient age (≥ 50 vs < 50 years), clinical tumor size (≥ 20 vs < 20 mm), nodal status (≥ 4 vs < 4 positive nodes), grade, treatment arm, and menopausal status, to evaluate the independence of CTX-prognostic from established clinicopathologic features. To evaluate CTX-benefit’s predictive ability of everolimus benefit, a Cox model including an interaction term between everolimus treatment and CTX-benefit score was fitted, with a significant interaction indicating differential everolimus effect with CTX-benefit score. Hazard ratios for continuous scores are reported per 1 SD increase in score.

### CTX Spatial Score Maps

Patch-level CTX-prognostic and CTX-benefit spatial score maps were generated for 30 patients selected from the extremes of the score distributions, spanning all recurrence risk and chemotherapy benefit categories. Two board-certified pathologists with subspecialty expertise in breast pathology, blinded to clinical outcomes, independently reviewed spatial score heatmaps alongside the corresponding H&E image to identify dominant histological features of each phenotype.

### Ethics Statement

All patients provided informed consent for participation in the UNIRAD trial. As this study is retrospective, no further consent was required.

## RESULTS

### CTX-prognostic stratifies patients by recurrence risk

To evaluate the prognostic performance of CTX-prognostic, we used 556 patients from the phase III UNIRAD trial (**Table 1**). CTX demonstrated good discriminative performance in predicting DFS, with a C-index of 0.67 (95% CI = 0.61-0.73). Among the 556 patients, 268\ (48%) were classified as high-risk, and 288 (52%) as low-risk. A total of 69 events were observed at a median follow-up of 4.97 years, of which 62 occurred within the first 5 years (KM-estimated cumulative incidence = 13.6%). When stratified by risk score, DFS differed significantly (log-rank p < 0.001) between the low- and high-risk groups, with a 5-year DFS of 80% (95% CI = 74%-85%) in high-risk patients compared with 93% (95% CI = 88%-95%) in low-risk patients (**Figure 2a**). A univariable Cox proportional hazards model found CTX-prognostic to be significantly associated with DFS (HR = 1.49, 95% CI = 1.26-1.77, Wald test p < 0.001, **Table S1**).

**FIG 2.**
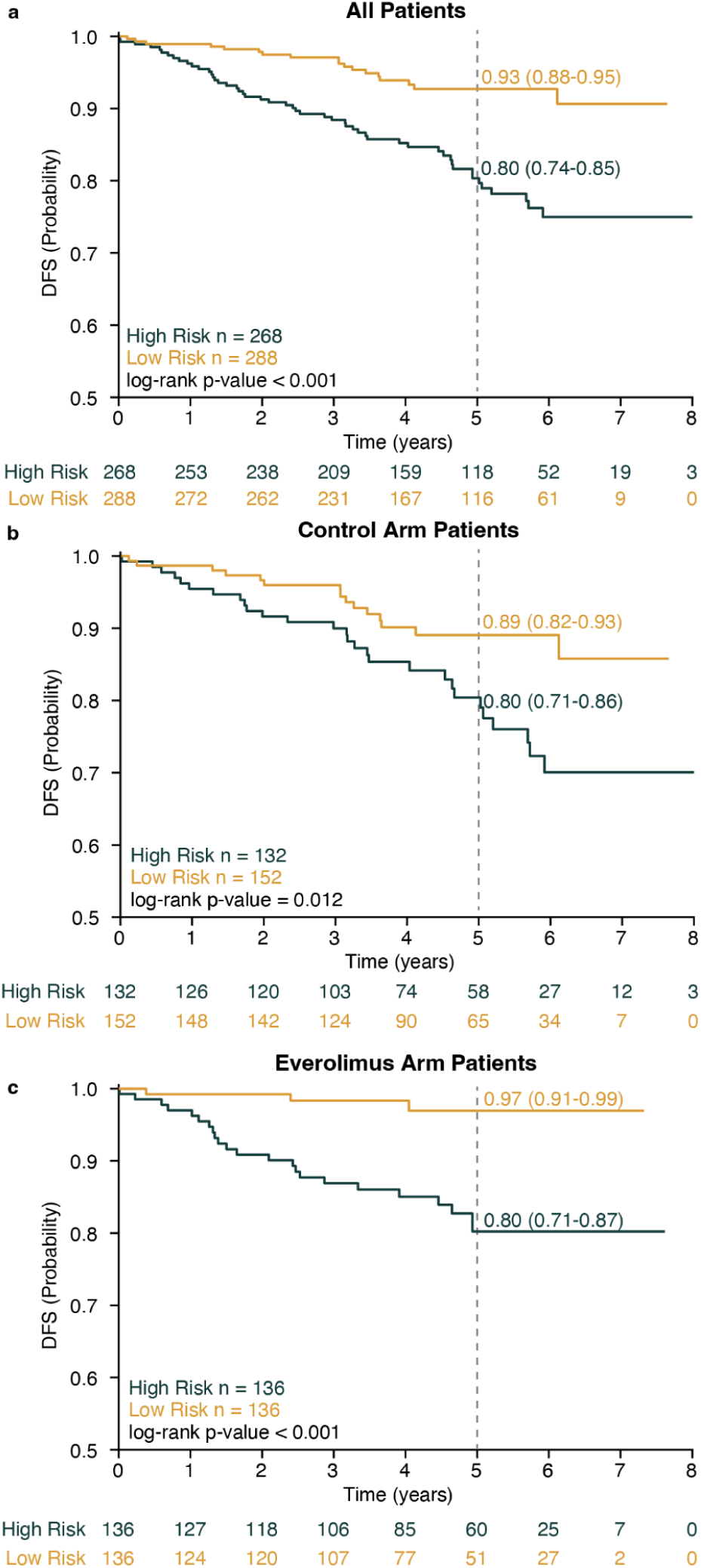
CTX demonstrated prognostic value in patients across treatment arms in the UNIRAD trial. a) KM estimates of probability of disease-free survival (DFS) stratified by CTX risk group across all patients, with 5-year DFS probabilities and corresponding 95% confidence intervals shown; p-value was derived from a two-sided log-rank test. Numbers below the curves represent the number of patients at risk at each time point. b) KM estimates of DFS probability in patients randomized to the control arm. c) KM estimates of DFS probability in patients randomized to the everolimus arm. Separation between risk groups was observed in the pooled cohort and within each arm separately, consistent with prognostic value independent of treatment received.

Because the multi-modal AI model incorporates both morphologic features from H&E-stained slides and clinical data, we evaluated whether the AI model captured prognostic information beyond clinical variables alone. In a multivariable Cox model, CTX-prognostic was the only variable that remained statistically significant (HR = 1.31, 95% CI = 1.06-1.61, Wald test p = 0.012, **Table S2**) after adjusting for treatment arm, tumor size, nodal status, age, grade, and menopausal status. A subset of patients (n = 454) had additional pathological variables available including lymphovascular invasion, ER positivity, PR positivity, HER2 positivity, lobular histology, and ductal histology. We performed an exploratory analysis adjusting for these additional covariates within this patient subset, finding that CTX-prognostic remained significantly prognostic for DFS (HR = 1.39, 95% CI = 1.07-1.81, Wald test p = 0.012, **Table S3**). Treatment arm was also significantly associated with DFS (HR = 0.53, 95% CI = 0.29-0.96, p = 0.04).

We next sought to determine whether CTX-prognostic stratifies recurrence risk within trial arms. Among patients randomized to the control arm, CTX had a C-index of 0.58 (95% CI = 0.49-0.66), indicating modest risk stratification. Patients classified as high-risk (5-year DFS = 80%, 95% CI = 71%-86%) had significantly lower DFS (log-rank p = 0.012) than low-risk patients (5-year DFS = 89%, 95% CI = 82%-93%, **Figure 2b**). A univariable Cox model found CTX-prognostic to be significantly associated with DFS (HR = 1.30, 95% CI = 1.03-1.65, Wald test p = 0.027, **Table S4**), though it did not remain significantly prognostic for DFS in multivariable analysis (HR = 1.11, 95% CI = 0.83-1.48, Wald test p = 0.49, **Table S5**). In patients randomized to receive everolimus, CTX-prognostic had a C-index of 0.59 (95% CI = 0.46-0.73). High-risk patients had significantly lower DFS (5-year DFS = 80%, 95% CI = 71%-87%) than patients classified as low-risk (5-year DFS = 97%, 95% CI = 91%-99%, log-rank p < 0.001, **Figure 2c**). A univariable Cox proportional hazards model also found that CTX-prognostic was significantly prognostic for DFS in patients treated with chemoendocrine therapy and everolimus (HR = 1.84, 95% CI = 1.42-2.39, p < 0.001, **Table S6**), and remained significant in a multivariable Cox model after adjusting for clinical features (HR = 1.69, 95% CI = 1.23-2.31, Wald test p = 0.001, **Table S7**).

### CTX stratifies risk across endpoints

We hypothesized that CTX would also be prognostic for the secondary endpoints, DMFS and OS. Indeed, when modeled as a continuous variable in a multivariable Cox model, CTX was associated with both DMFS (HR = 1.27, 95% CI = 1.02-1.58, Wald test p = 0.036, **Table S8**) and OS (HR = 1.63, 95% CI = 1.20-2.21, Wald test p = 0.002, **Table S9**). The C-index was 0.65 (95% CI = 0.58-0.72), 0.71 (95% CI = 0.57-0.83) for DMFS, and 0.71 (95% CI = 0.57-0.83) OS, respectively. Using the pre-selected cut-off, patients classified as high-risk had a lower DMFS (5-year DMFS = 82%, 95% CI = 77%-87%, **Figure S2a**) and OS (5-year OS = 92%, 95% CI = 88%-95%, **Figure S2b**) than low-risk patients (5-year DMFS: 93%, 95% CI = 89%-96%, log-rank p < 0.001; 5-year OS = 98%, 95% CI = 95%-99%, log-rank p < 0.001).

### CTX-benefit stratifies patients by everolimus benefit

We next examined whether CTX-benefit identified patients who may benefit from everolimus. We hypothesized that patients at high risk of recurrence but with low responsiveness to adjuvant chemotherapy may benefit from treatment escalation with agents such as everolimus. To test this, we compared DFS between the everolimus and control arms within each CTX-benefit category. Among patients predicted to benefit from chemotherapy, 5-year DFS was 84% (95% CI = 70%-90%) in the control arm and 70% (95% CI = 55%-81%) in the everolimus arm, with an observed HR of 1.29 (95% CI = 0.64-2.61, log-rank p = 0.481, **Figure 3a**). These data neither support nor exclude an everolimus effect in this subgroup with statistical significance. In contrast, among patients predicted not to benefit from chemotherapy, everolimus significantly improved DFS (log-rank p = 0.005), with a 5-year DFS of 85% (95% CI = 79%-90%) in the control arm compared to 95% (95% CI = 90%-97%) in the everolimus arm (**Figure 3b**).

**FIG 3.**
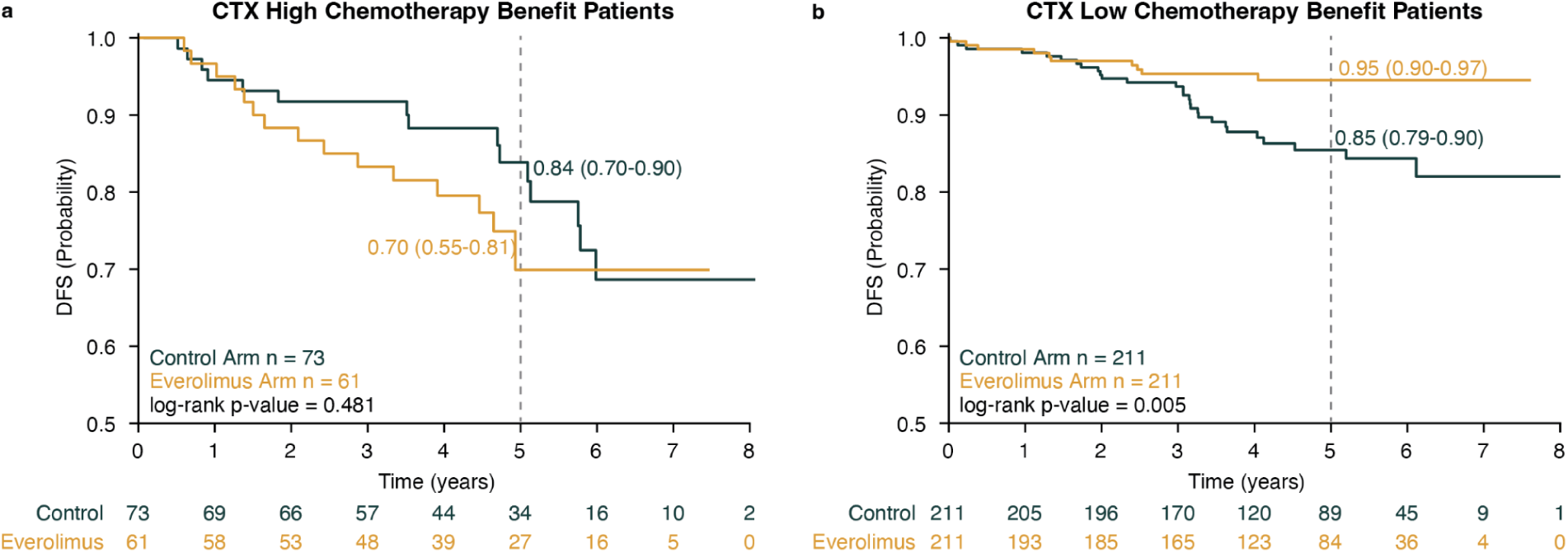
Kaplan-Meier estimates of 5-year DFS by CTX chemotherapy benefit group, comparing the control arm with the everolimus arm. KM plots of patients in the a) CTX high-benefit group and b) CTX low-benefit group, with patients randomized to the control arm in green and patients in the everolimus arm in yellow. Statistical significance was evaluated with a two-sided log-rank test. Numbers displayed on the plots indicate 5-year disease-free survival probability estimates with corresponding 95% confidence intervals. Numbers below represent the number of patients at risk at each time point.

To determine if CTX-benefit was a predictor of everolimus benefit, we fit a Cox proportional hazards model including an interaction term between CTX-benefit score and everolimus treatment. From this model, we found that the interaction between CTX-benefit score and everolimus treatment was statistically significant (HR = 1.57, 95% CI = 1.08-2.29, Wald test p = 0.02). Next, to assess whether this interaction was independent of established clinical factors, we fit a multivariable Cox model adjusting for tumor size, nodal status, age at diagnosis, grade, and menopausal status (**Table 2**). The interaction term (HR = 1.68, 95% CI = 1.12-2.53, Wald test p = 0.01), treatment arm (p = 0.01), and tumor size (p = 0.04) were statistically significant.

**TABLE 2:** Hazard ratios and interaction term from a Cox proportional hazards model for DFS, including a CTX-benefit by everolimus treatment interaction term. The interaction term was statistically significant, indicating that the association between everolimus treatment and DFS differed by CTX-benefit score.

| **Variable** | **HR** | **Lower 95% CI** | **Upper 95% CI** | **p-value** |
| --- | --- | --- | --- | --- |
| **CTX-benefit** | 1.05 | 0.78 | 1.41 | 0.74 |
| **Everolimus treatment** | 0.49 | 0.28 | 0.86 | 0.01 |
| **CTX-benefit x treatment** | 1.68 | 1.12 | 2.53 | 0.01 |
| **Tumor size** | 2.24 | 1.04 | 4.80 | 0.04 |
| **Nodal status** | 1.63 | 0.96 | 2.75 | 0.07 |
| **Age** | 1.10 | 0.58 | 2.07 | 0.77 |
| **Grade** | 1.01 | 0.64 | 1.58 | 0.97 |
| **Menopausal status** | 1.00 | 0.62 | 1.61 | 0.99 |

### Histological correlates of CTX risk and benefit phenotypes

To investigate the biological basis of CTX stratification, we evaluated CTX-prognostic and CTX-benefit spatial score heatmaps (**Figure 4**). The low-risk/low-benefit phenotype included cases with well-differentiated, often mucinous tumors with prominent in situ disease and a low volume of invasive disease. The high-risk/high-benefit phenotype was characterized by high cellularity, prominent mitotic activity, infiltrative cords of tumor cells, pockets of lymphocytic inflammation, fat involvement, and poorly differentiated morphology. The high-risk/low-benefit phenotype was distinguished by prominent in situ disease, intermediate cellularity, and mucin, with invasive carcinoma occasionally approaching the skin surface.

**FIG 4.**
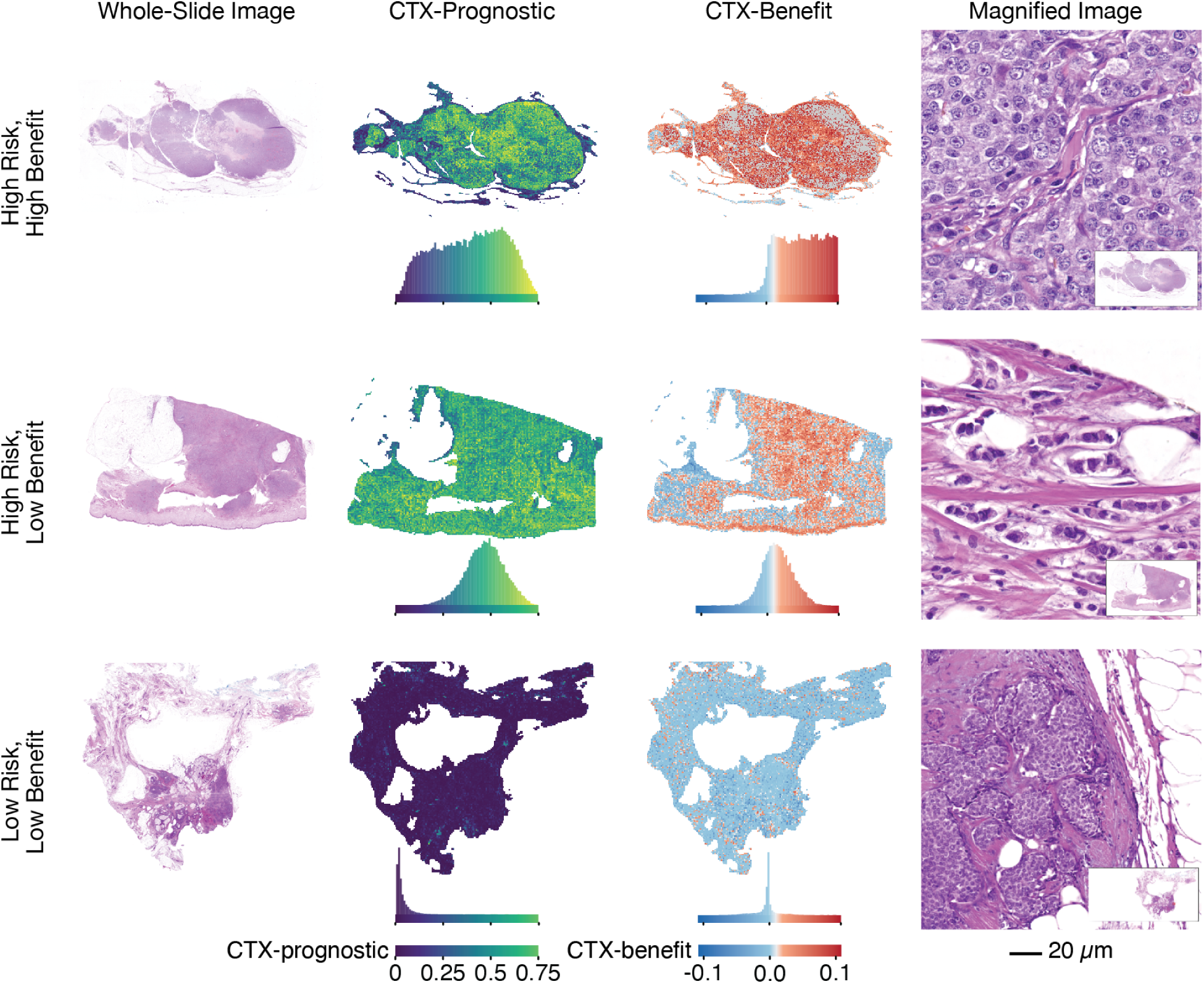
Histological correlates of CTX prognostic and benefit predictions in three representative patients. Each row shows one patient, selected to illustrate the high-risk/high-benefit (top), high-risk/low-benefit (middle), and low-risk/low-benefit (bottom) phenotypes. The first column from the left shows the H&E whole-slide image. The spatial risk map, where CTX-prognostic score was calculated for each WSI patch individually (dark purple = low CTX-prognostic, green = high CTX-prognostic), is in the second column. The third column displays the spatial chemotherapy-benefit map, where CTX-benefit score was calculated for each WSI patch individually (blue = low CTX-benefit, red = high CTX-benefit). The final column displays a magnified H&E field with inset. Histograms show per-patch score distributions. Scale bar, 20 µm.

## DISCUSSION

In this study, we demonstrate that Ataraxis Breast CTX provides prognostic and predictive information in the phase III UNIRAD trial, which enrolled a cohort of high-risk early breast cancer patients. CTX-prognostic stratified patients by recurrence risk and identified a subgroup of node-positive patients whose 5-year DFS outcomes exceeded 90% despite meeting clinical high-risk criteria. CTX-benefit, developed to predict chemotherapy benefit, identified patients who derived benefit from escalation therapy with everolimus independent of established clinicopathologic features.

This finding addresses an urgent clinical need, as current escalation strategies are applied to broadly defined risk groups and no validated biomarker identifies which patients derive benefit.^19^ Recent trials of treatment escalation in early HR+/HER2− breast cancer have demonstrated modest absolute improvements in invasive disease-free survival. In the phase III NATALEE trial, adjuvant ribociclib demonstrated a 4.5% absolute improvement in 5-year iDFS (85.5% vs 81.0% with endocrine therapy alone),^6^ and the phase III monarchE trial showed a 7.6% absolute increase with abemaciclib (83.6% vs 76.0%).^20^ CTX low-risk patients in UNIRAD achieved a 5-year DFS rate of 93% across randomized arms despite meeting high-risk eligibility criteria. As all patients in the UNIRAD trial would be eligible for adjuvant ribociclib, these data suggest CTX may identify a subgroup with sufficiently low residual risk that further escalation with additional systemic agents may offer limited absolute benefit, leading to overtreatment. Notably, while a prior biomarker analysis of the UNIRAD cohort also suggested the presence of a low-risk subgroup, that study excluded everolimus-treated patients, survival differences between UNIRAD risk groups did not reach statistical significance, and did not evaluate prediction of everolimus benefit.^21^

The phase II TAMRAD and phase III BOLERO-2 trials established that adding everolimus to endocrine therapy in patients with metastatic HR+/HER2− breast cancer increased progression-free survival.^22,23^ However, its efficacy in early-stage disease is limited. The UNIRAD and SWOG 1207 trials both failed to show improvement in disease-free survival. Additionally, everolimus tolerability was poor, as more than half of patients discontinued treatment before completing the planned course due to adverse events.^7^ Within this context, our findings suggest that predictive biomarkers such as CTX-benefit may enable enrichment for patients most likely to derive benefit, potentially reconciling negative trial results with heterogeneity in treatment sensitivity.

High-risk patients predicted by CTX to benefit from chemotherapy may be more sensitive to cytotoxic therapy, whereas those with little or no predicted chemotherapy benefit may retain greater residual risk despite receiving adjuvant chemotherapy. Therefore, these patients may represent a subgroup that is more likely to derive benefit from targeted therapies such as everolimus. Consistent with this, we observed in our cohort that a subset of high-risk patients with in situ disease and intermediate cellularity had improved outcomes with everolimus, suggesting that distinct biological features may underlie sensitivity to chemotherapy versus endocrine therapy combined with targeted therapy. Therefore, CTX may support a strategy in which escalation beyond standard adjuvant therapies is reserved for patients with high residual risk who are unlikely to derive benefit from chemotherapy.

This analysis has limitations. First, the high rate of treatment discontinuation due to toxicity from everolimus may have influenced the observed benefit. Second, patients who received neoadjuvant chemotherapy were excluded, and therefore CTX’s performance in this patient population is unknown. Finally, only 56% of patients enrolled in the UNIRAD trial had H&E slides available, and patients with and without slides available differed modestly in T stage, nodal status, grade, and menopausal status (standardized mean differences: 0.17-0.23). Further validation in an external cohort, such as SWOG 1207, may confirm generalizability of our results.

Collectively, our findings indicate that Ataraxis Breast CTX provides prognostic risk stratification and predicts benefit from adjuvant everolimus in patients with clinically high-risk HR+/HER2− breast cancer. By identifying patients with minimal residual risk following chemoendocrine therapy, CTX may establish a patient population that would derive little to no absolute improvement in DFS from additional therapies. The CTX-benefit score suggested that clinically high-risk patients who are unlikely to benefit from adjuvant chemotherapy may be the best candidates for further adjuvant therapy escalation. More broadly, these results highlight the potential of causal, multi-modal modeling to refine therapeutic decision-making and move from clinical variable-based treatment-intensification decisions toward personalized, AI-enhanced care.

## SUPPLEMENTAL FIGURES

**FIG S1.**
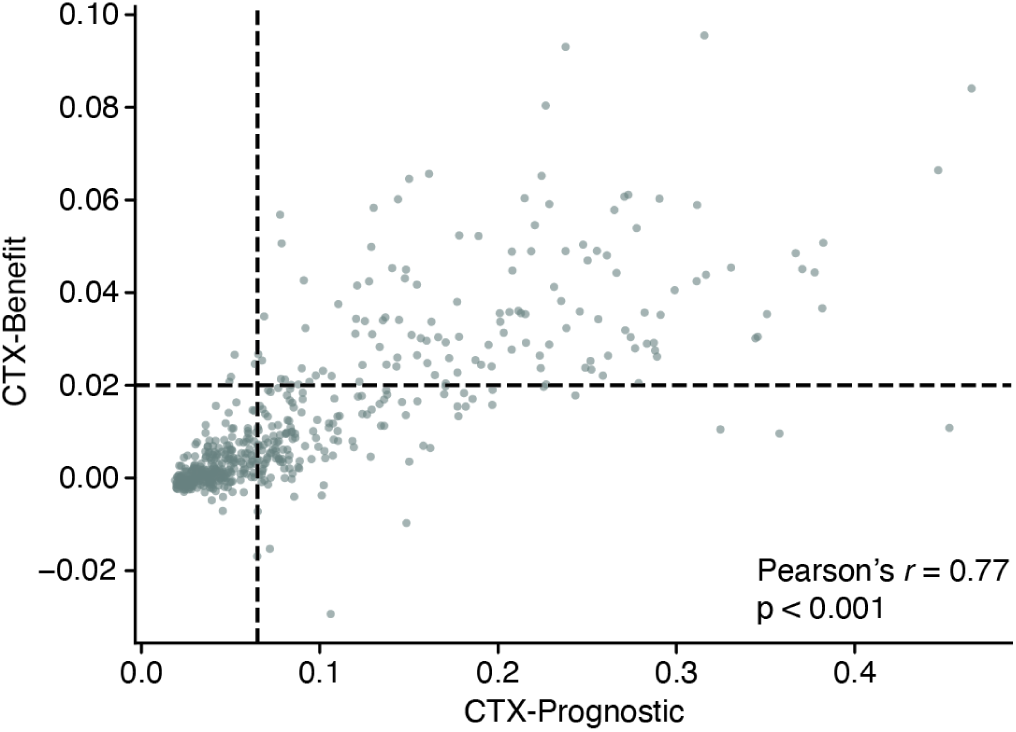
Comparison of CTX-prognostic with CTX-benefit. Scatterplot of CTX-prognostic scores and CTX-benefit scores. Patients in the bottom-right quadrant (n = 138) are patients who were predicted to have high risk of recurrence but derive little chemotherapy benefit. Four patients classified as low-risk were predicted to benefit from chemotherapy. CTX-prognostic and CTX-benefit scores were correlated but not redundant.

**FIG S2.**
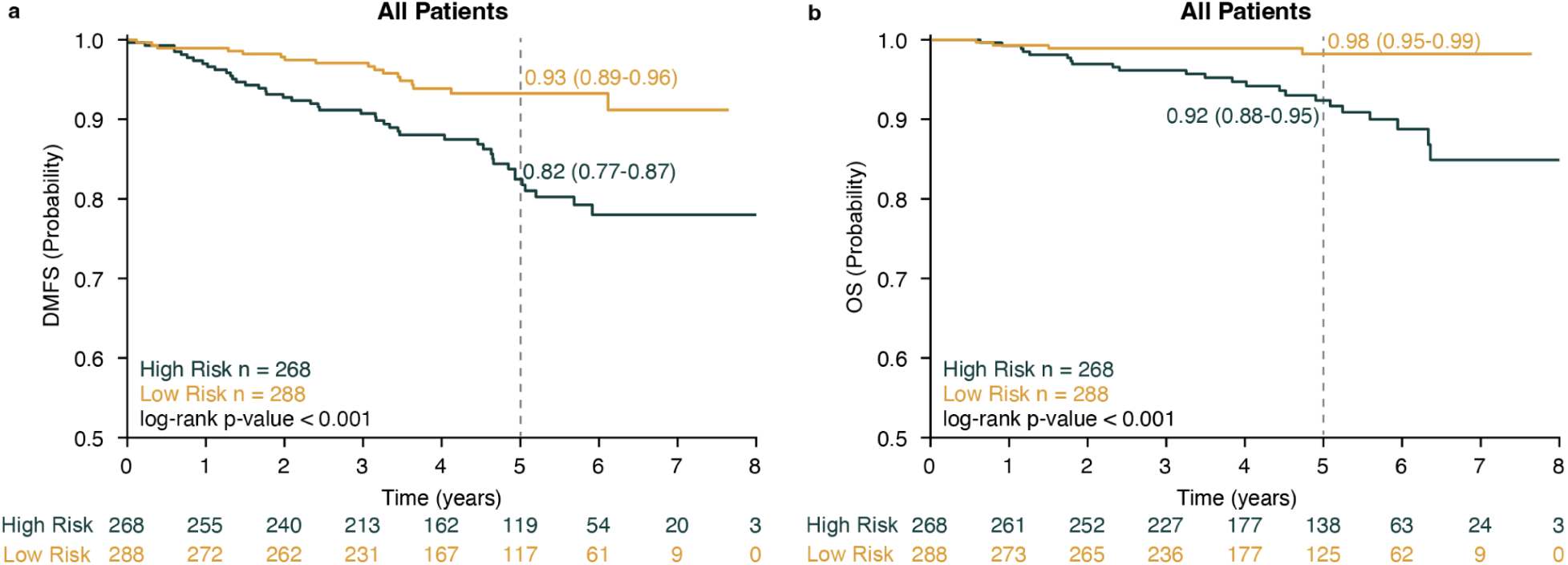
CTX stratifies patients by secondary endpoints. KM estimates of probability of a) distant metastasis-free survival (DMFS) and b) overall survival (OS) stratified by CTX-prognostic risk group across all patients. Five-year group-wise estimates and corresponding 95% confidence intervals shown; p-values were derived from a two-sided log-rank test. Numbers below represent the number of patients at risk at each time point.

## SUPPLEMENTAL TABLES

**TABLE S1:**
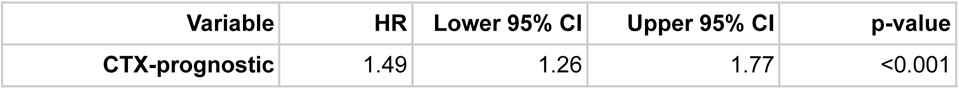
Hazard ratios from a Cox proportional hazards regression model evaluating the association of CTX-prognostic score with DFS (n = 556). CTX-prognostic was significantly prognostic with DFS.

**TABLE S2:** Hazard ratios from a Cox proportional hazards regression model evaluating the association of CTX-prognostic score, clinical variables, and treatment arm with DFS (n = 556). CTX-prognostic remained significantly prognostic for DFS after adjustment for clinical variables.

| Variable | HR | Lower 95% CI | Upper 95% CI | p-value |
| --- | --- | --- | --- | --- |
| **CTX-prognostic** | 1.31 | 1.06 | 1.61 | 0.01 |
| **Treatment arm** | 0.66 | 0.41 | 1.08 | 0.10 |
| **Tumor size** | 2.06 | 0.96 | 4.40 | 0.06 |
| **Nodal status** | 1.46 | 0.83 | 2.57 | 0.18 |
| **Age** | 1.03 | 0.54 | 1.93 | 0.94 |
| **Grade** | 1.17 | 0.76 | 1.78 | 0.47 |
| **Menopausal status** | 0.94 | 0.57 | 1.53 | 0.79 |

**TABLE S3:** Hazard ratios from a Cox proportional hazards regression model evaluating the association of CTX-prognostic score, clinical variables, treatment arm, and additional pathological variables with DFS (n = 454). CTX-prognostic remained significantly associated with DFS after adjustment in this exploratory analysis.

| **Variable** | **HR** | **Lower 95% CI** | **Upper 95% CI** | **p-value** |
| --- | --- | --- | --- | --- |
| **CTX-prognostic** | 1.39 | 1.07 | 1.81 | 0.01 |
| **Treatment arm** | 0.53 | 0.29 | 0.96 | 0.04 |
| **Tumor size** | 2.13 | 0.82 | 5.54 | 0.12 |
| **Nodal status** | 1.46 | 0.74 | 2.88 | 0.27 |
| **Age** | 0.87 | 0.41 | 1.82 | 0.70 |
| **Grade** | 1.56 | 0.91 | 2.68 | 0.17 |
| **Menopausal status** | 0.85 | 0.48 | 1.51 | 0.56 |
| **LVI** | 0.93 | 0.52 | 1.67 | 0.82 |
| **ER positivity** | 1.00 | 0.98 | 1.02 | 0.93 |
| **PR positivity** | 1.00 | 0.99 | 1.01 | 0.61 |
| **HER2 positivity** | 0.99 | 0.98 | 1.01 | 0.41 |
| **Ductal histology** | 1.10 | 0.40 | 3.03 | 0.85 |
| **Lobular histology** | 1.39 | 0.55 | 3.55 | 0.49 |

**TABLE S4:**
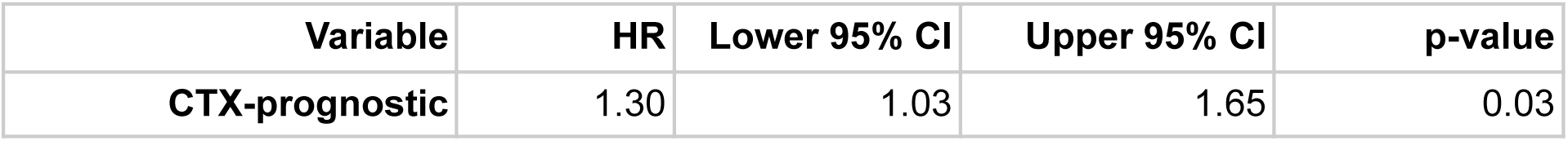
Hazard ratios from a univariable Cox proportional hazards regression model evaluating the association of CTX-prognostic with DFS for patients in the control arm (n = 284). CTX-prognostic was significant in patients randomized to the control arm.

**TABLE S5:** Hazard ratios from a Cox proportional hazards regression model evaluating the association of CTX-prognostic score and clinical covariates with DFS for patients in the control arm (n = 284). CTX-prognostic was not significant in patients randomized to the control arm after adjustment.

| Variable | HR | Lower 95% CI | Upper 95% CI | p-value |
| --- | --- | --- | --- | --- |
| **CTX-prognostic** | 1.11 | 0.83 | 1.48 | 0.49 |
| **Tumor size** | 1.67 | 0.70 | 3.94 | 0.25 |
| **Nodal status** | 1.73 | 0.85 | 3.51 | 0.13 |
| **Age** | 0.87 | 0.39 | 1.92 | 0.73 |
| **Grade** | 0.95 | 0.55 | 1.63 | 0.85 |
| **Menopausal status** | 0.92 | 0.50 | 1.70 | 0.80 |

**TABLE S6:** Hazard ratios from a univariable Cox proportional hazards regression model evaluating the association of CTX-prognostic with DFS for patients in the everolimus arm (n = 272). CTX-prognostic was significant in patients treated with everolimus.

| Variable | HR | Lower 95% CI | Upper 95% CI | p-value |
| --- | --- | --- | --- | --- |
| **CTX-prognostic** | 1.84 | 1.42 | 2.39 | <0.001 |

**TABLE S7:** Hazard ratios from a Cox proportional hazards regression model evaluating the association of CTX-prognostic score and clinical covariates with DFS for patients in the everolimus arm (n = 272). CTX-prognostic was significant in patients treated with everolimus after adjustment.

| Variable | HR | Lower 95% CI | Upper 95% CI | p-value |
| --- | --- | --- | --- | --- |
| **CTX-prognostic** | 1.69 | 1.23 | 2.31 | 0.001 |
| **Tumor size** | 5.93 | 0.79 | 44.19 | 0.08 |
| **Nodal status** | 1.14 | 0.44 | 2.90 | 0.79 |
| **Age** | 1.57 | 0.52 | 4.76 | 0.43 |
| **Grade** | 1.74 | 0.85 | 3.54 | 0.13 |
| **Menopausal status** | 1.22 | 0.51 | 2.90 | 0.66 |

**TABLE S8:**
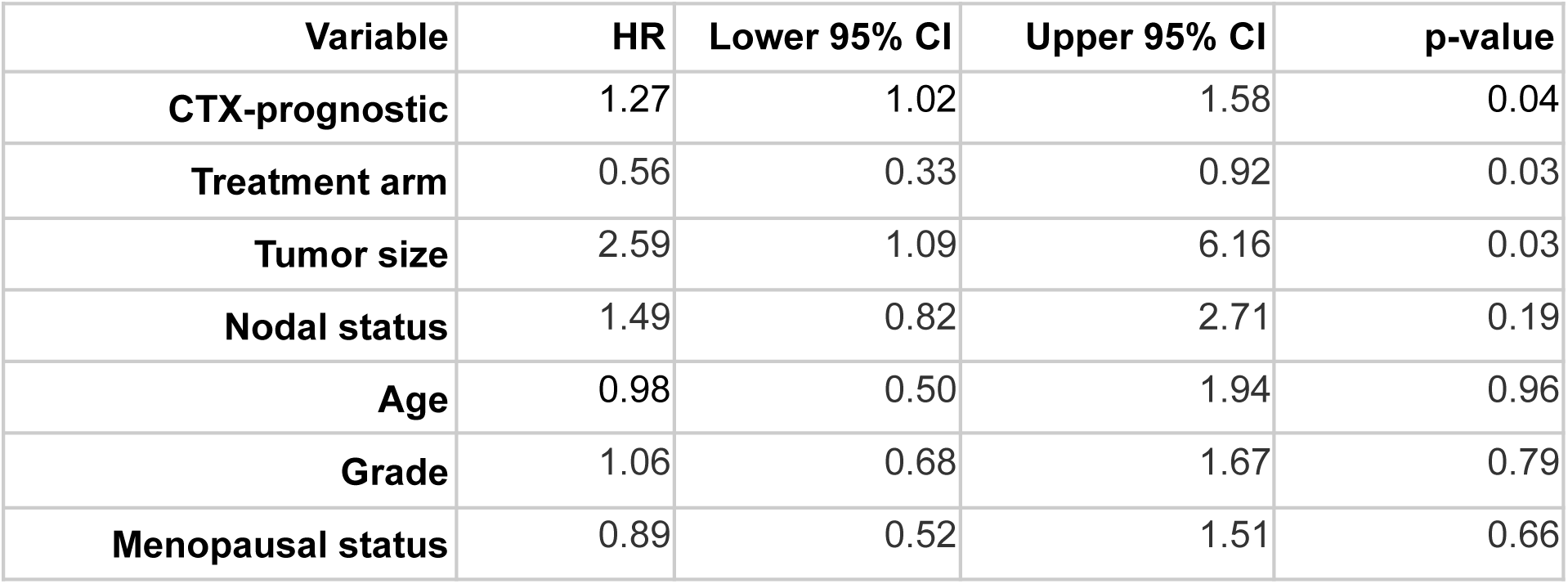
Hazard ratios from a Cox proportional hazards regression model evaluating the association of CTX-prognostic score, clinical covariates, and treatment arm with DMFS. CTX-prognostic was significantly prognostic for DMFS.

| **Variable** | **HR** | **Lower 95% CI** | **Upper 95% CI** | **p-value** |
| --- | --- | --- | --- | --- |
| **CTX-prognostic** | 1.27 | 1.02 | 1.58 | 0.04 |
| **Treatment arm** | 0.56 | 0.33 | 0.92 | 0.03 |
| **Tumor size** | 2.59 | 1.09 | 6.16 | 0.03 |
| **Nodal status** | 1.49 | 0.82 | 2.71 | 0.19 |
| **Age** | 0.98 | 0.50 | 1.94 | 0.96 |
| **Grade** | 1.06 | 0.68 | 1.67 | 0.79 |
| **Menopausal status** | 0.89 | 0.52 | 1.51 | 0.66 |

**TABLE S9:**
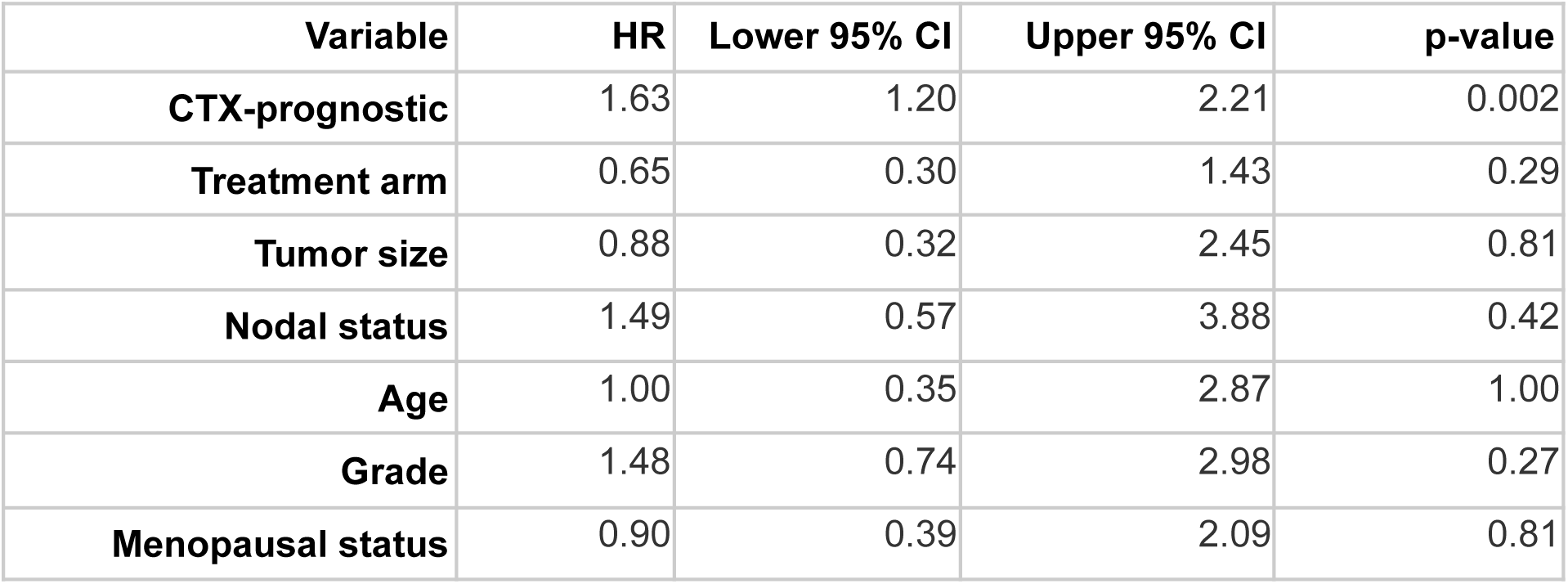
Hazard ratios from a Cox proportional hazards regression model evaluating the association of CTX-prognostic score, clinical covariates, and treatment arm with OS. CTX-prognostic was significantly prognostic for OS.

## ACKNOWLEDGEMENTS

The authors thank the patients whose data were included in the UNIRAD cohort, along with the institutions that made the data available for research.

## DATA AVAILABILITY

The data is not publicly available due to institutional and ethical constraints. Access can be requested directly from Unicancer.

## CODE AVAILABILITY

Ataraxis Breast CTX is available for non-commercial research use upon reasonable request. A Jupyter notebook to reproduce the analyses presented in this study is available upon request.

